# Racial disparities in nephrectomy and mortality among patients with renal cell carcinoma: Findings from SEER

**DOI:** 10.1101/2022.11.02.22281850

**Authors:** Joshua Ikuemonisan, Taiwo Opeyemi Aremu, Isaac Oyejinmi, Christopher Ajala, Nnabuchi Anikpezie, Oyindamola Akinso, Mutsa Mtengwa, Adeyemo David, Olugbenga Olokede, Oluwakayode Adejoro

## Abstract

**Purpose:** To assess racial differences in the receipt of nephrectomy in patients diagnosed RCC in the US.

**Materials and Methods:** 2005 to 2015 data from the SEER database was analyzed and 70059 patients with RCC were identified. We compared demographic and tumor characteristics between Blacks and Whites. We applied logistic regression to assess the influence of race on the odds of the receipt of nephrectomy. We also applied Cox proportional hazards model to assess the impact of race on cancer-specific mortality (CSM) and all-cause mortality (ACM) in patients diagnosed with RCC in the US.

**Results:** Overall, there was a relative increase in the use of nephrectomy from 2007 (p<0.0001). However, Blacks had 18% lower odds of receiving nephrectomy compared to Whites (p < 0.0001). The odds of the receipt of nephrectomy also reduced with age at diagnosis. In addition, patients with T3 stage had the greatest odds of receiving nephrectomy when compared to T1 (p < 0.0001). There was no difference in the risk of cancer-specific mortality between Blacks and Whites, Blacks had 27% greater odds of all-cause mortality than Whites (p < 0.0001). Patients who did not receive nephrectomy had a 42% and 35% higher risk of CSM and ACM respectively, when compared to patients who received nephrectomy.

**Conclusions:** Blacks diagnosed with RCC in the US have a greater ACM risk and are less likely than Whites to receive nephrectomy. Systemic changes are needed to eliminate racial disparity in the treatment and outcomes of RCC in the US.

## Introduction

Kidney cancer is a form of cancer that occurs in the tissues of the kidney. The main types of kidney cancer are renal cell cancer (RCC), transitional cell cancer, and Wilms tumor [1], with RCC being the most common type of kidney cancer and constituting approximately 90% of cancers derived from the renal parenchyma [2-5]. Kidney cancer is the 8th most common cancer in the United States (US) [6,7]. This is a disease of public health importance; in the United States, the prevalence of kidney and renal pelvis cancer was 582,727 as of 2018. In 2021 alone, the estimated number of new kidney cancer cases in the US was 76,080, accounting for about 4.0% of all new cancer cases in the US in 2021[7]. Kidney cancer is one of the top ten cancers that lead to death, accounting for 2.3% of all estimated cancer deaths in the US in 2021[7].

Although kidney cancer impacts persons of all races, studies have shown that it does not always affect the races equally [8]. Literature is replete with evidence of racial disparities in health conditions and outcomes between Blacks and Whites in the US and how the experiences of different races have shaped health outcomes [9]. Additionally, the incidence rates of kidney cancers have been rising more rapidly among Blacks than Whites over the past five decades [10]. An analysis of the incidence and survival among patients with kidney cancer from the national Surveillance, Epidemiology, and End Results (SEER) Registry Database, showed that Blacks have a more significant rise in incidence and a poorer outcome than White patients of the same age and disease stage [11].

Non-metastatic kidney cancers are treated with nephrectomy. While this procedure is not without risks, evidence suggests that Black are more likely to experience worse treatment outcomes than their white counterparts. In addition, Blacks have more renal-relevant comorbidities (hypertension, diabetes, and chronic renal failure) and poorer overall cancer survival compared to Whites [12,13].

Studies evaluating disparity in nephrectomy by race have been few and many have yielded equivocal results, necessitating the investigation of the role of race in the receipt of nephrectomy using a large and nationally representative database. Therefore, this study examines the presentation, early definitive surgical treatment, and mortality among patients with RCC in the US, to investigate racial disparity in the receipt of nephrectomy for renal cell carcinoma patients.

## Methods

### Data Source

We analyzed SEER data of patients diagnosed with renal cell carcinoma between 2005 and 2015 in the United States. The SEER database is a product of a nationally representative surveillance conducted by the National Cancer Institute (NCI), which collects demographic, pathologic, and clinical information on incident cancer across 18 geographic locations in the United States, covering nearly a third of the US population. Our study cohort includes patients diagnosed with renal cell carcinoma between January 1, 2005, and December 31, 2015 [14].

### Study Population

We identified 75,714 patients diagnosed with histologically confirmed non-metastatic renal cell carcinoma diagnosed between 2005 and 2015. We excluded patients who were diagnosed for the first time in nursing/ convalescent homes (n=242), patients whose race were neither Black nor White (n=5,029) as the size of the patients from other racial groups were small compared to that of Whites or Blacks, and patients diagnosed in Louisiana in 2005 (n=384). The records from patients diagnosed with RCC in Louisiana were excluded because of the possibility of a differential loss of records due to the impact of hurricane Katrina in 2005. The final cohort consisted of 70,059 patients with renal cell carcinoma. The outcomes of the study were reviewed until December 31, 2015, or the patient’s demise.

### Ethical Consideration

This study analyzed publicly available data from the SEER database, hence has been approved based on the broad agreement signed in the SEER data-use agreement. This agreement ensures the study satisfies the Health Insurance Portability and Accountability Act of 1996 (HIPAA), confidentiality, and ethical considerations.

### Variables

The baseline demographic data include age (10–year intervals), race (Black vs White), tumor stage (T1, T2, T3, and T4), receipt of nephrectomy (Yes vs No), binary survival status (Dead (Coded as “0”) vs Alive (Coded as “1”)), and SEER registry geographic location according to the Census Bureau of the United States (Northeast, Midwest, South, and West). We defined survival as the number of months following diagnosis of RCC to the earlier of death or the date of the end of the study. The patients in this database who received nephrectomy had their surgery within 4– 6 months of diagnosis. The variables were operationalized as categorical variables except for “year of diagnosis” and “survival” variables that were operationalized as continuous variables.

### Outcomes

The unit of analysis was the patient. The primary outcome was the receipt of nephrectomy, which was either radical or partial nephrectomy. CSM (cancer-specific mortality) and ACM (all-cause mortality) were the secondary outcome variables operationalized as time-to-event variables.

### Analysis

Chi-squared and T-test univariate analyses were used to compare baseline characteristics of Black vs White patients in the study. The influence of race on receipt of nephrectomy was evaluated by fitting a multivariable logistic regression model, with demographic and tumor characteristics as covariates. Cox proportional hazard models were fitted to determine the influence of race on CSM and ACM while controlling for confounders. Additionally, Cox proportional hazard models were fitted to assess the relationship between nephrectomy and survival in the overall study population and within each race category. All analyses were conducted using SAS 9.4 software (Cary, NC). The level of statistical significance was set at *p<* 0.05.

## Results

Black and White patients constituted 11.51% (n=8,066) and 88.49% (n=61,993) of the study cohort respectively. The post-diagnosis follow-up of the cohort median (interquartile range) was 54 (63) months; the median (interquartile range) post-diagnosis follow-up for Black and White patients was 51 (60) months and 54 (63) months, respectively. The mean age at diagnosis (SD) was 61.4 (12.4) years; the mean (SD) for Blacks and Whites was 59.5 (11.7) years and 61.7 (12.5) years, respectively. Figure 1 summarizes the trend of the use of nephrectomy. There was a relative increase in the use of nephrectomy among patients diagnosed from 2007 through 2014 with a slight decline in 2015 (p < 0.0001).

Table 1 details the comparison of demographic and tumor characteristics of all study subjects, and Black versus White patients. Approximately 57.8% (n=40,484) were ≤ 65 years, 74.4% (n=52,151) had T1 disease. There were significant differences across tumor stages and geographical location of patients by race (p < 0.0001). A lower proportion of Black patients received nephrectomy compared to Whites (82.8% vs 85.4%; p < 0.0001). A greater proportion of White vs Black patients died of RCC (11.7% vs 9.1 %, p < 0.0001) but no difference in all cause death between Whites vs Blacks (25.8% vs 26.3%, p = 0.34).

**Table 1.** Characteristic of study cohort and comparison by race

| Covariates | Total (70059: 100%) | Black 8066 (11.51%) | White 61993 (88.49%) | p value |
| --- | --- | --- | --- | --- |
| year of diagnosis |  |  |  |  |
| 2005 | 4809 (6.86) | 466 (5.78) | 4343 (7.01) | <.0001 |
| 2006 | 5667 (8.09) | 543 (6.73) | 5124 (8.27) |  |
| 2007 | 6095 (8.70) | 679 (8.42) | 5416 (8.74) |  |
| 2008 | 6582 (9.39) | 777 (9.63) | 5805 (9.36) |  |
| 2009 | 6903 (9.85) | 803 (9.96) | 6100 (9.84) |  |
| 2010 | 6294 (8.98) | 734 (9.10) | 5560 (8.97) |  |
| 2011 | 6355 (9.07) | 788 (9.77) | 5567 (8.98) |  |
| 2012 | 6499 (9.28) | 777 (9.63) | 5722 (9.23) |  |
| 2013 | 6556 (9.36) | 802 (9.94) | 5754 (9.28) |  |
| 2014 | 6962 (9.94) | 827 (10.25) | 6135 (9.90) |  |
| 2015 | 7337 (10.47) | 870 (10.79) | 6467 (10.43) |  |
| Age at diagnosis, years |  |  |  |  |
| <45 | 6565 (9.37) | 831 (10.30) | 5734 (9.25) | <.0001 |
| 45 - 54 | 13168 (18.80) | 1760 (21.82) | 11408 (18.40) |  |
| 55 - 64 | 20751 (29.62) | 2656 (32.93) | 18095 (29.19) |  |
| 65 - 74 | 18946 (27.04) | 2033 (25.20) | 16913 (27.28) |  |
| 75 - 84 | 9324 (13.31) | 726 (9.00) | 8598 (13.87) |  |
| ≥85 | 1305 (1.86) | 60 (0.74) | 1245 (2.01) |  |
| Regions |  |  |  |  |
| Midwest | 8470 (12.09) | 1219 (15.11) | 7251 (11.70) | <.0001 |
| Northeast | 10009 (14.29) | 1077 (13.35) | 8932 (14.41) |  |
| South | 18354 (26.20) | 3519 (43.63) | 14835 (23.93) |  |
| West | 33226 (47.43) | 2251 (27.91) | 30975 (49.97) |  |
| Tumor stage |  |  |  |  |
| T1 | 52151 (74.44) | 6598 (81.80) | 45553 (73.48) | <.0001 |
| T2 | 298 (0.43) | 53 (0.66) | 245 (0.40) |  |
| T3 | 16451 (23.48) | 1303 (16.15) | 15148 (24.44) |  |
| T4 | 1159 (1.65) | 112 (1.39) | 1047 (1.69) |  |
| Tumor grade |  |  |  |  |
| 1 | 8996 (12.84) | 1022 (12.67) | 7974 (12.86) | <.0001 |
| 2 | 36588 (52.22) | 4175 (51.76) | 32413 (52.28) |  |
| 3 | 19554 (27.91) | 2406 (29.83) | 17148 (27.66) |  |
| 4 | 4921 (7.02) | 463 (5.74) | 4458 (7.19) |  |
| Nephrectomy |  |  |  |  |
| No | 10439 (14.90) | 1384 (17.16) | 9055 (14.61) | <.0001 |
| Yes | 59620 (85.10) | 6682 (82.84) | 52938 (85.39) |  |
| Partial<br>Nephrectomy |  |  |  |  |
| No | 46230 (65.99) | 5272 (65.36) | 40958 (66.07) | 0.2068 |
| Yes | 23829 (34.01) | 2794 (34.64) | 21035 (33.93) |  |
| Radical<br>Nephrectomy |  |  |  |  |
| No | 34268 (48.91) | 4178 (51.80) | 30090 (48.54) | <.0001 |
| Yes | 35791 (51.09) | 3888 (48.20) | 31903 (51.46) |  |
| Cancer-specific<br>mortality |  |  |  |  |
| Alive | 62047 (88.56) | 7331 (90.89) | 54716 (88.26) | <.0001 |
| Dead | 8012 (11.44) | 735 (9.11) | 7277 (11.74) |  |
| All-cause mortality |  |  |  |  |
| Alive | 51946 (74.15) | 5945 (73.70) | 46001 (74.20) | 0.3355 |
| Dead | 18113 (25.85) | 2121 (26.30) | 15992 (25.80) |  |

Table 2 summarizes the comparison of demographic and tumor characteristics of all study subjects by their nephrectomy status. Approximately 85.1% (n = 59,620) of the patients received nephrectomy. Table 2 presents differences in nephrectomy by age, sex, race, region, and tumor stage. Patients younger than 65 years were more likely to receive nephrectomy than older patients diagnosed with RCC (< 0.0001), and patients with lower-stage disease were more likely to receive nephrectomy (p < 0.0001).

**Table 2.** Characteristics of the study cohort and comparison by receipt of nephrectomy

| Covariates | Total (70059; 100%) | Yes (59620; 85.10%) | No (9541; 13.62%) | p value |
| --- | --- | --- | --- | --- |
| *Year of diagnosis* |  |  |  |  |
| 2005 | 4809 (6.86) | 4118 (6.91) | 691 (6.62) | 0.0001 |
| 2006 | 5667 (8.09) | 4772 (8.00) | 895 (8.57) |  |
| 2007 | 6095 (8.70) | 5087 (8.53) | 1008 (9.66) |  |
| 2008 | 6582 (9.39) | 5519 (9.26) | 1063 (10.18) |  |
| 2009 | 6903 (9.85) | 5830 (9.78) | 1073 (10.28) |  |
| 2010 | 6294 (8.98) | 5337 (8.95) | 957 (9.17) |  |
| 2011 | 6355 (9.07) | 5440 (9.12) | 915 (8.77) |  |
| 2012 | 6499 (9.28) | 5571 (9.34) | 928 (8.89) |  |
| 2013 | 6556 (9.36) | 5658 (9.49) | (8.60) |  |
| 2014 | 6962 (9.94) | 6007 (10.08) | 955 (9.15) |  |
| 2015 | 7337 (10.47) | 6281 (10.54) | 1056 (10.12) |  |
| *Age at diagnosis,*<br>*years* |  |  |  |  |
| <45 | 6565 (9.37) | 5852 (9.82) | 713 (6.83) | 0.0001 |
| 45 - 54 | 13168 (18.80) | 11525 (19.33) | 1643 (15.74) |  |
| 55 - 64 | 20751 (29.62) | 17941 (30.09) | 2810 (26.92) |  |
| 65 - 74 | 18946 (27.04) | 16016 (26.86) | 2930 (28.07) |  |
| 75 - 84 | 9324 (13.31) | 7375 (12.37) | 1949 (18.67) |  |
| ≥85 | 1305 (1.86) | 911 (1.53) | 394 (3.77) |  |
| Regions |  |  |  |  |
| Midwest | 8470 (12.09) | 6505 (10.91) | 1965 (18.82) | 0.0001 |
| Northeast | 10009 (14.29) | 8440 (14.16) | 1569 (15.03) |  |
| South | 18354 (26.20) | 15795 (26.49) | 2559 (24.51) |  |
| West | 33226 (47.43) | 28880 (48.44) | 4346 (41.63) |  |
| Tumor stage |  |  |  |  |
| T1 | 52151 (74.44) | 44406 (74.48) | 7745 (74.19) | 0.0001 |
| T2 | 298 (0.43) | 138 (0.23) | 160 (1.53) |  |
| T3 | 16451 (23.48) | 14396 (24.15) | 2055 (19.69) |  |
| T4 | 1159 (1.65) | 680 (1.14) | 479 (4.59) |  |
| Tumor grade |  |  |  |  |
| 1 | 8996 (12.84) | 7249 (12.16) | 1747 (16.74) | 0.0001 |
| 2 | 36588 (52.22) | 31324 (52.54) | 5264 (50.43) |  |
| 3 | 19554 (27.91) | 16848 (28.26) | 2706 (25.92) |  |
| 4 | 4921 (7.02) | 4199 (7.04) | 722 (6.92) |  |
| Cancer-specific mortality |  |  |  |  |
| Alive | 62047 (88.56) | 53276 (89.36) | 8771 (84.02) | 0.0001 |
| Dead | 8012 (11.44) | 6344 (10.64) | 1668 (15.98) |  |
| All-cause mortality |  |  |  |  |
| Alive | 51946 (74.15) | 45238 (75.88) | 6708 (64.26) | 0.0001 |
| Dead | 18113 (25.85) | 14382 (24.12) | 3731 (35.74) |  |
| Race |  |  |  |  |
| Black | 8066 (11.51) | 6682 (11.21) | 1384 (13.26) | 0.0001 |
| White | 61993 (88.49) | 52938 (88.79) | 9055 (86.74) |  |

Table 3 displays the results of the multivariable logistic regression. The odds of receipt of nephrectomy reduced with increasing age. Compared to Whites, Blacks had 18% lower odds of receipt of nephrectomy. Compared to patients with T1, patients with stages T2, T3, T4 disease were 83% less likely, 17% more likely, and 78% less likely to receive nephrectomy, respectively.

**Table 3.** Multivariable logistic regression model showing the odds of receipt of nephrectomy

| Covariates | HR (95% CI) | p value |
| --- | --- | --- |
| *Year of diagnosis* | 1.01 (1.01 – 1.02) | 0.0006 |
| *Age at diagnosis, years* |  |  |
| <45 |  |  |
| 45 - 54 | 0.86 (0.78 - 0.95) | 0.0017 |
| 55 - 64 | 0.79 (0.71 – 0.85) | <.0001 |
| 65 - 74 | 0.65 (0.60 - 0.71) | <.0001 |
| 75 - 84 | 0.46 (0.41 – 0.50) | <.0001 |
| ≥85 | 0.27 (0.24 – 0.31) | <.0001 |
| *Race* |  |  |
| White (reference) |  |  |
| Black | 0.82 (0.76- 0.87) | <.0001 |
| *Region* |  |  |
| Midwest (reference) |  |  |
| Northeast | 1.68 (1.56 - 1.81) | <.0001 |
| South | 1.87 (1.75 - 2.00) | <.0001 |
| West | 2.03 (1.94 - 2.16) | <.0001 |
| *Tumor stage* |  |  |
| T1 (reference) |  |  |
| T2 | 0.17 (0.13 - 0.21) | <.0001 |
| T3 | 1.17 (1.11 - 1.24) | <.0001 |
| T4 | 0.22 (0.19 - 0.25) | <.0001 |
| *Tumor grade* |  |  |
| 1 (reference) |  |  |
| 2 | 1.41 (1.33 - 1.50) | <.0001 |
| 3 | 1.55 (1.44 - 1.66) | <.0001 |
| 4 | 1.60 (1.43 - 1.77) | <.0001 |

Table 4 summarizes the mortality risks of the study patients. The risk of CSM is similar between Blacks and Whites. However, CSM increased with older age and higher tumor stage. Patients who did not receive nephrectomy had a 42% higher risk of CSM relative to patients who received nephrectomy. The risk of ACM in Blacks was 27% higher compared to Whites. ACM increased with older age and higher stage. Patients who did not receive nephrectomy had a 35% higher risk of ACM relative to patients who received nephrectomy. The risk of CSM and ACM of the study patients, by race, showed a similar trend in patients treated with either radical or partial nephrectomy (Table 5A & 5B).

**Table 5A.** Multivariable Cox proportional hazards models showing the risk of mortality in the radical nephrectomy study cohort

| Covariates | Cancer-specific mortality |  | All-cause mortality |  |
| --- | --- | --- | --- | --- |
|  | HR (95% CI) | p value | HR (95% CI) | p value |
| *Race* |  |  |  |  |
| White (reference) |  |  |  |  |
| Black | 1.08 (1.00 - 1.17) | 0.0497 | 1.29 (1.23 - 1.35) | <.0001 |

**Table 5B.** Multivariable Cox proportional hazards models showing the risk of mortality in the partial nephrectomy study cohort

| Covariates | Cancer-specific mortality |  | All-cause mortality |  |
| --- | --- | --- | --- | --- |
|  | HR (95% CI) | p value | HR (95% CI) | p value |
| *Race* |  |  |  |  |
| White (reference) |  |  |  |  |
| Black | 1.09 (1.01 - 1.18) | 0.0317 | 1.29 (1.23 - 1.35) | <.0001 |

## Discussion

In this study, we investigated the difference in the receipt of nephrectomy and mortality risk between Black and White patients, diagnosed with renal cell carcinoma in the US over ten years. Blacks had lower odds of receipt of nephrectomy and a higher risk of all-cause mortality than Whites. In agreement with our findings, a study by Zini et al. suggests that the receipt of nephrectomy, the gold standard treatment for non-metastatic renal cell cancer, differs by race. However, it suggests that this difference did not have any effect on survival outcomes [15]. Another study examining the trends in the use of nephrectomy and cytoreductive surgery in the US reported that Blacks were less likely than Whites to receive surgical intervention when diagnosed with RCC [16].

The use of nephrectomy appears to be on the rise, and most patients in this study received some form of nephrectomy. Although younger age at diagnosis was associated with greater odds of receipt of nephrectomy and Blacks were more likely to be diagnosed with RCC at a younger age than Whites, Blacks still had 18% lesser odds of nephrectomy receipt than Whites in this study. The SEER programs did not collect information on comorbidity or information that may help to calculate comorbidity scores (e.g., Charlosn Comorbidty Index). Hence the influence of comorbidity on receipt of nephrectomy could not be determined. Previous study examining the influence of age and comorbidity of receipt of radical surgeries for major urological cancers suggest age is more important than comorbidity in the receipt of radical surgeries [17]. While age and comorbidity alone may not be the only factors limiting RCC health outcomes in Blacks, poor access and utilization of health care, distrust for the health system and a lack of health education are likely contributors to poorer health outcomes in Blacks.

Blacks had higher ACM than Whites when diagnosed with renal cell carcinoma in this study. Increasing age at diagnosis, tumor grade, and stage are associated with greater odds of cancer-specific and all-cause mortality. Because Blacks have 18% lesser odds of receipt of nephrectomy than Whites, it is not surprising that Blacks have about 27% greater odds of ACM than Whites. Multiple factors such as differential comorbidities and use of other therapies such as adjuvant therapy not captured in this analysis due to the limitation of the data could have affected the ACM outcome. A study that analyzed SEER data concluded that there is reduced receipt of nephrectomy in Blacks versus Whites but suggests that the reduction in receipt of nephrectomy does not influence survival outcomes. The study had a smaller population, a shorter follow-up time and due to its timing, was unlikely to have captured the benefits of nephron-sparing surgery [15].

Access to nephrectomy and survival outcomes are lower in Blacks than in Whites [15, 18]. Compared to Whites, Blacks are less likely to be insured, have an established source of healthcare, and experience greater difficulty in seeking healthcare [19-21]. Also, there is a long-standing distrust for the healthcare system among Blacks [22,23]. Perhaps, the most discussed example of this is the Tuskegee syphilis experiment, where Blacks were intentionally exposed to harm. Similar to this is the experience of African American women unethically subjected to gynecological procedures and such experiences as when slaves were not offered medical care because they were slaves [21]. Similarly, it is reported that providers caring for Blacks are less likely to give adequate health information to their patients, thus impairing their ability to participate in shared decision-making about their care [24]. These experiences may have bred a fear of complications, an aversion to surgery [25,26] and an overall negative attitude towards accepting procedures such as nephrectomy. The discrimination in health care delivery plus the negative predisposition towards the health care system contributes to poorer health outcomes in Blacks diagnosed with RCC.

### Limitations

We applied a retrospective study design. Due to the nature of the SEER data, it was impossible to capture the use of additional therapies such as adjuvant therapies/ chemotherapies which may have effects on survival outcomes. Lastly, information on comorbidities was not available in our data, hence, we were unable to adjust for comorbidities in our analysis.

## Conclusion

Although there has been technological advancement and the use of nephrectomy in treating patients diagnosed with RCC has improved, Blacks are still less likely than Whites to receive nephrectomy despite the greater likelihood of RCC incidence and mortality in Blacks. Several factors stemming from systemic racism and racial inequalities contribute greatly to a disparity in health care delivery. Improving health outcomes for patients diagnosed with RCC will require a deliberate adoption of system-wide social and medical changes in the approach to health care delivery and treatment decisions that address systemic racism and eliminate disparity in health outcomes in Blacks and other minority populations in the US.

## Data Availability

The data presented in this study are available on request from the corresponding author.

## Key abbreviations

ACM: (All-cause mortality)
CSM: (Cancer-specific mortality)
RCC: (Renal cell carcinoma)
SEER: (Surveillance, Epidemiology, and End Results)

